# Investigation of factors associated with delayed initiation of breastfeeding in Papua New Guinea: a cross-sectional study

**DOI:** 10.1101/2024.09.18.24313839

**Authors:** McKenzie Maviso, Francis Pulsan, Lisa M. Vallely

**Affiliations:** Division of Public Health, School of Medicine and Health Sciences, University of Papua New Guinea, Port Moresby, Papua New Guinea; Division of Clinical Sciences, School of Medicine and Health Sciences, University of Papua New Guinea, Port Moresby, Papua New Guinea; Asia and Pacific Health Program, Kirby Institute, UNSW Sydney, Australia

## Abstract

**Background:** Breastfeeding within the first hour of birth is critical for newborn survival. However, in Papua New Guinea (PNG), about 40% of newborns are not breastfed within the first hour of birth. This study aimed to determine the prevalence and factors associated with delayed breastfeeding initiation in PNG.

**Methods:** This study utilised secondary data from the 2016–2018 PNG Demographic and Health Survey (DHS), a nationally representative cross-sectional study. A total weighted sample of 4748 women aged 15–49 were included. Complex samples analysis was performed to determine the direction of association between the independent variables and delayed initiation of breastfeeding.

**Results:** About a quarter (24.6%) of women delayed initiation of breastfeeding. Women with an unplanned pregnancy (AOR 1.32; 95% CI: 1.03 to 1.68), those who had a caesarean section (AOR 3.16; 95% CI: 1.39 to 7.17), those who did not initiate newborn skin-to-skin contact immediately after birth (AOR 1.83; 95% CI: 1.41 to 2.38), and those who watched television (AOR 1.39; 95% CI: 1.11 to 1.75), or were from the Momase region (AOR 1.31; 95% CI: 0.88 to 1.93) had higher odds of delayed breastfeeding initiation. Conversely, those who read a newspaper or magazine (AOR 0.76; 95% CI: 0.61 to 0.95), were from the Southern (AOR 0.81; 95% CI: 0.56 to 1.15) and Highlands (AOR 0.86; 95% CI: 0.58 to 1.29) regions, and gave birth at home or in the village (AOR 0.69; 95% CI: 0.49 to 0.96) had lower odds of delayed initiation of breastfeeding.

**Conclusion:** One in four women in this study delayed initiation of breastfeeding until after one hour after birth. Interventions to promote optimal breastfeeding require a multi-sectoral approach, as well as bolstering health workers’ capacity to encourage and support early initiation of breastfeeding during the antenatal and early postnatal periods.

**What is known about the subject:**

- Early breastfeeding initiation within the first hour of birth is an important intervention to reduce childhood morbidities and mortalities.
- In PNG, 40% of newborns are not breastfed immediately or within the first hour of birth.

**What this study adds:**

- One in four women in PNG delayed early initiation of breastfeeding.
- Unplanned pregnancy, caesarean section, no newborn skin-to-skin contact immediately after birth, and exposure to media predicted delayed initiation of breastfeeding.
- Birth at home or in the village reduces the likelihood of delayed initiation of breastfeeding.

**How this study affects research, practice or policy:**

- This study seeks to promote targeted breastfeeding support for mothers, particularly in rural areas.
- Breastfeeding advocacy and promotion programmes, strengthening health workers’ capacity to address factors influencing delayed initiation of breastfeeding, and promoting optimal breastfeeding practices are required.

## Introduction

Scaling up breastfeeding is one of the most cost-effective interventions for improving infant and child survival and reducing under-five mortality.^1^ Breastfeeding has the potential to save the lives of over 800 000 children under the age of five every year.^2^ It is universally accepted and recommended that all newborn babies, irrespective of the place of birth, be placed in skin-to-skin contact with their mothers immediately after birth to promote breastfeeding initiation within the first hour of birth.^3^ Early initiation of breastfeeding is the provision of the mother’s breast milk within the first hour of birth and ensures that the newborn receives colostrum, while delayed breastfeeding initiation occurs after one hour of birth.^3,4^

Early initiation of breastfeeding has health benefits for both the infant and the mother. It provides many nutritional, immunological, and psychosocial benefits, including protection of the infant against infectious diseases and enhanced maternal-infant bonding.^3^ For the mothers, early initiation of breastfeeding promotes the release of oxytocin for uterine contractions to prevent postpartum haemorrhage.^5,6^ In addition, initiation of breastfeeding within the first hour and breastfeeding exclusively for up to six months has been shown to lower the odds of cancer, diabetes (Type 2), and cardiovascular diseases, including maternal mortality.^7–9^

Despite the established health benefits of early initiation of breastfeeding, a higher rate of suboptimal breastfeeding remains in low- and middle-income countries (LMICs).^4,10^ There is also increasing evidence that delayed initiation of breastfeeding (after the first hour of birth) has been associated with neonatal infections and illnesses,^4,7,11^ and increased newborn and child mortality.^7,11^ Findings from a systematic review revealed that initiating breastfeeding between 2 and 23 hours after birth increased the risk of neonatal death by 33% compared to breastfeeding within the first hour of birth.^11^ The delay in initiating breastfeeding was also found to increase the risk of respiratory diseases, including breathing difficulty during the first 6 months.^11^ This can influence breastfeeding and negatively affect its continuation into infancy.

In Papua New Guinea (PNG), the rate of early initiation of breastfeeding is around 60%,^12^ suggesting that 40% of newborns are at higher risk of morbidity and mortality throughout their infancy and early childhood years. Earlier work from PNG has identified low rates of early initiation of breastfeeding are associated with factors such as younger maternal age, lack of breastfeeding knowledge and support, lack of antenatal education, place and mode of birth, grandmothers’ involvement, and socio-cultural norms.^12–14^ While there are few studies in PNG on infant feeding practices^14–16^ including early breastfeeding initiation,^12^ there is a paucity of information concerning factors predisposing to delayed initiation of breastfeeding. Understanding the factors that influence the delay in initiating breastfeeding is crucial for designing interventions to improve breastfeeding outcomes in line with the country’s *Infant and Young Child Feeding Policy.*^17^ Therefore, this study aimed to determine the prevalence and factors associated with delayed initiation of breastfeeding in PNG.

## Methods

### Study design and data source

This study used secondary data from the 2016–2018 PNG Demographic and Health Survey (DHS), a nationally representative cross-sectional survey that collected demographic and health-related data across the country’s four major administrative regions: Southern, Highlands, Momase, and Islands.^18^ The current DHS was the second round of surveys conducted in the country by the National Statistical Office (NSO) with financial and technical assistance from Inner City Fund (ICF) International through The DHS Program, funded by the United States Agency for International Development (USAID). For sampling, the DHS used a two-stage stratified sampling procedure to sample census units from each stratum. In the first stage, 800 census units were within each sampling stratum to achieve implicit stratification and proportional allocation via a probability proportional-to-size selection. The second stage involved systematically selecting 24 households from each cluster using probability sampling, producing 19 200 households.^18^ In the interviewed households, 18 175 women aged 15–49 were identified for individual interviews, of which 15 198 women were interviewed successfully (a response rate of 84%). For this study, a weighted sample of 4748 women who had given birth three years before the survey and had complete information on the variables of interest was included. Information about the methodology, pretesting, sampling design, and selection is available in the final report^18^ and can be accessed online at https://dhsprogram.com/publications/publication-fr364-dhs-final-reports.cfm.

### Variable Measurement

#### Dependent variable

The dependent variable of interest was “delayed” initiation of breastfeeding. Information regarding the timing of initiation of breastfeeding was obtained from women using the question, “How long after birth did you first put (name of child) to the breast?” The dataset included four responses for breastfeeding initiation: “immediately,” “within the first hour,” “hours,” and “days.” A new variable for the time of initiation of breastfeeding was generated and dichotomized as “1” for “early” (immediately or within the first hour of birth), and “2” for “delayed” (any time after the first hour of birth).

#### Independent variables

The selection of the variables from potential confounders of delayed initiation of breastfeeding was identified in previous studies.^4,10,19^ Variable selection also depended on the availability of relevant data in the 2016–2018 PNGDHS. Potential individual and group-level factors associated with delayed initiation of breastfeeding are presented in the conceptual framework to guide the conceptualization (**figure 1**). The conceptual framework summarises selected independent variables at two levels that influenced the delay in breastfeeding initiation. Individual-level factors relate directly to the mother and infant and include maternal age, marital status, last pregnancy planned, education, occupation, place of residence and region. Infant characteristics include the infant’s sex, birth weight, birth size, birth order, newborn skin-to-skin contact immediately after birth, and prelacteal feeds. In addition, group-level factors are directly related to household and healthcare settings. Household characteristics include the husband’s education and occupation, wealth index, reading a newspaper or magazine, listening to the radio, and watching television, whereas health service characteristics include access to and utilisation of maternal and child health services, such as antenatal care visits, mode of birth, and place of birth (see **figure 1**).

**Figure 1.**
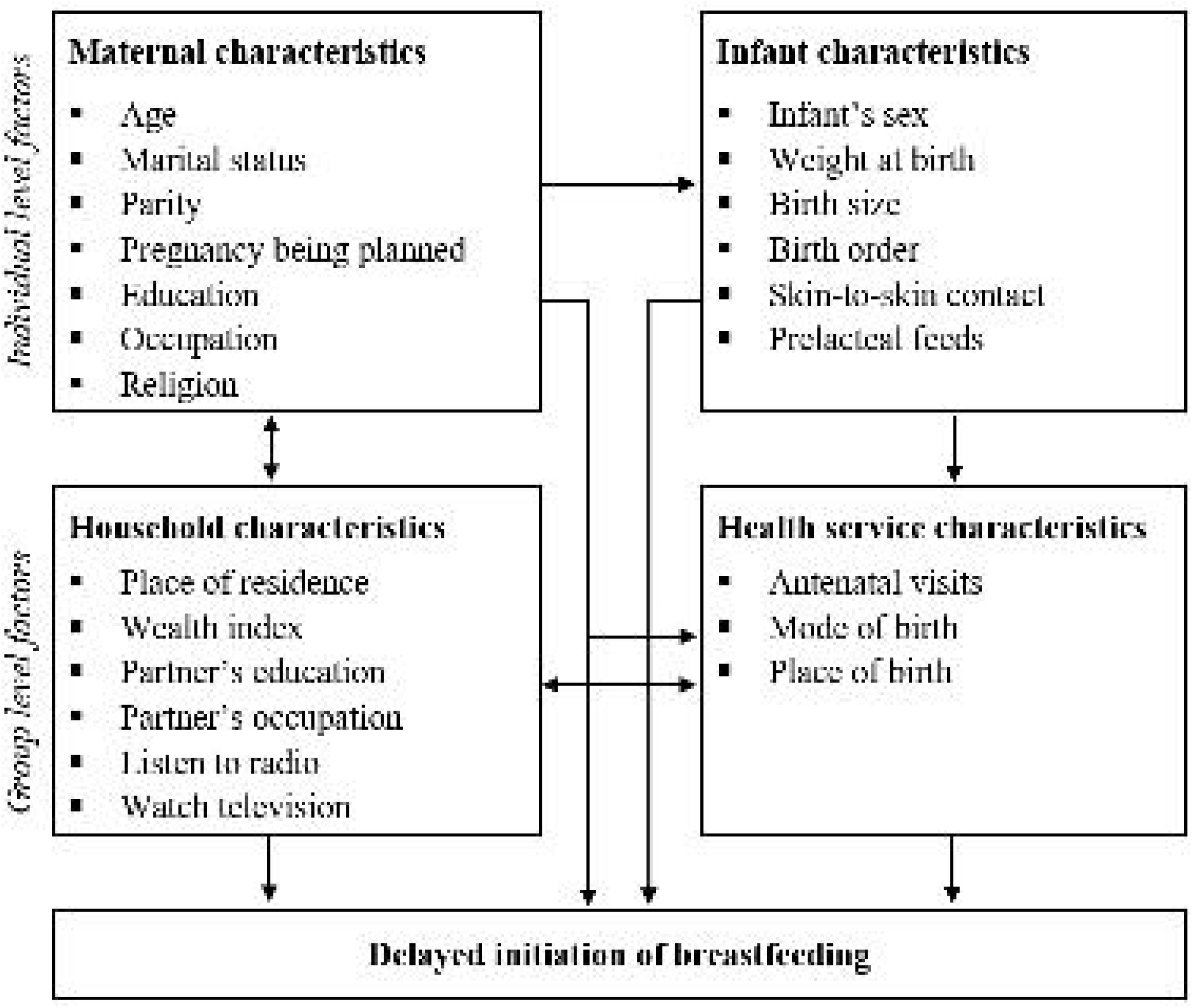
A conceptual framework of individual and group-level factors associated with delayed initiation of breastfeeding in PNG. Adapted from previous studies.^44,45^

#### Statistical analysis

The extracted data was cleaned and all the missing observations were dropped while subcategories of variables with small observations were merged. The final sample data was weighted to restore representativeness and produce a reliable estimate and standard error.

Descriptive statistics were computed and reported as weighted frequencies and percentages. Bivariate associations between predictors and delayed breastfeeding initiation were performed to determine the strength of association using Pearson’s chi-square test. Only significant variables from the bivariate analyses were entered into the multivariable logistic regression model. To account for the cluster sampling design and sample weight and provide generalizable and accurate estimates of proportion, probability values, and odds ratios,^20^ the complex samples analysis technique was applied. The Hosmer-Lemeshow goodness-of-fit was used to test for model fitness. Adjusted Odds Ratios (AORs) with 95% Confidence Intervals (CIs) are reported. A p≤0.05 was considered statistically significant. Data were analysed using IBM SPSS Statistics for Windows, Version 22.0 (IBM Corp., Armonk, NY, USA).

#### Patient and public involvement

Patients and/or the public were not involved in the design, conduct, reporting or dissemination of this research.

## Results

### Characteristics of the study population

**Table 1** presents the characteristics of the participants. A total weighted sample of 4748 women was included in the analysis. The mean (standard deviation) maternal age was 29.9 (±7.0) years. Overall, 1177 [24.6% (95% CI: 23 to 26)] of women delayed initiation of breastfeeding, which remained high in rural areas (88.3%). Half (50%) had attained primary education. More than two-thirds were not working (68.3%) and were living in rural areas (88.3%). Regarding maternal health factors, the majority of women had planned their last pregnancies (75.6%); more than half had received four or more antenatal visits (55.9%) and 61% gave birth in a health facility. In addition, over half of the women read newspapers/magazines (50.9%) and listened to radio (54.1%). Regarding infant-related factors, nearly all the newborns weighed over 2500g (81.4%) and 61.3% had no skin-to-skin contact immediately after birth. In Chi-square analysis, maternal age, last pregnancy planned, region of residence, husband’s education, wealth index, reading a newspaper or magazine, watching television, number of antenatal care visits, mode of birth, place of birth, birth size, birth order, newborn skin-to-skin contact, and prelacteal feeding were factors significantly associated with delayed breastfeeding initiation (p<0.05).

**Table 1.**
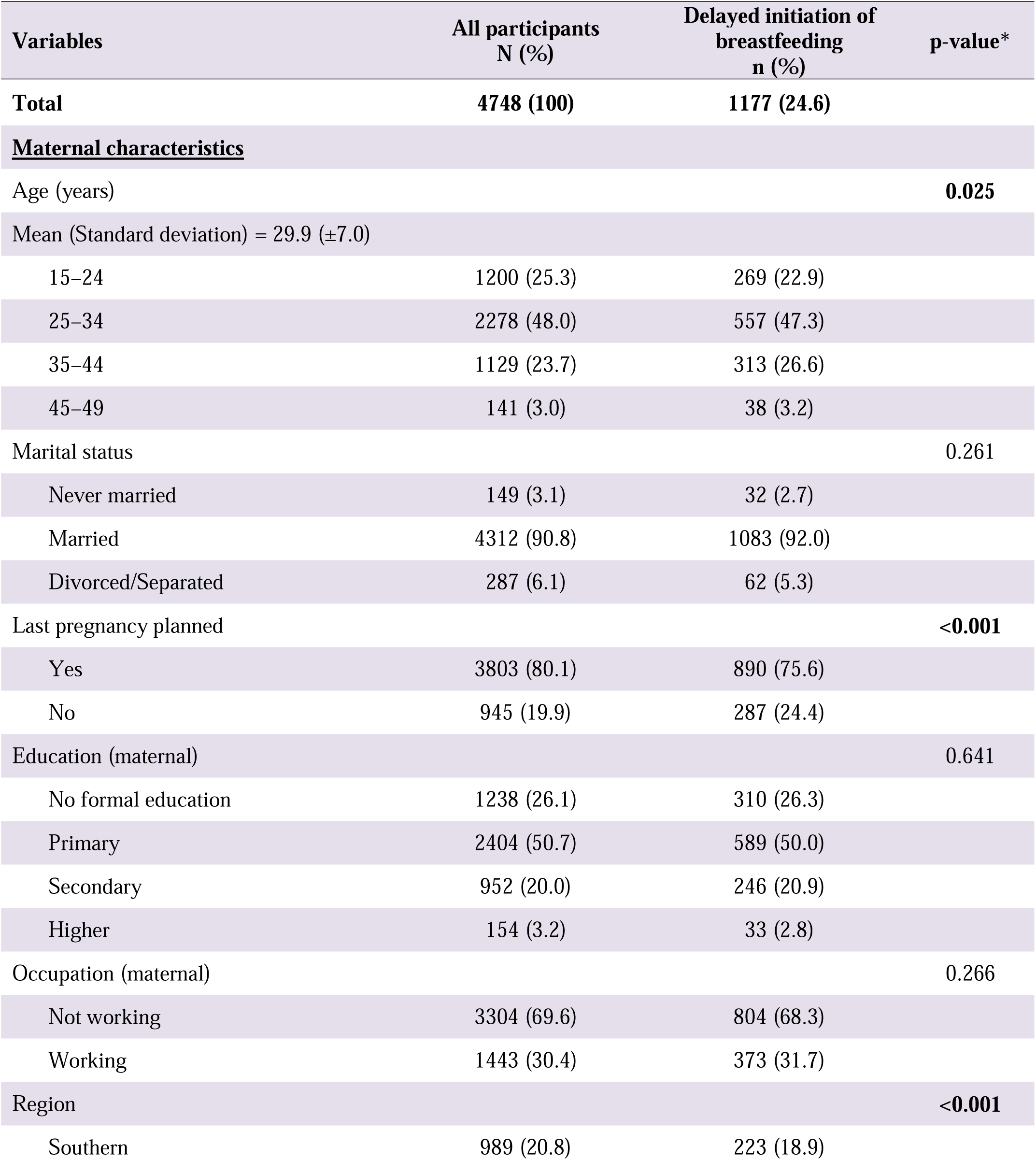
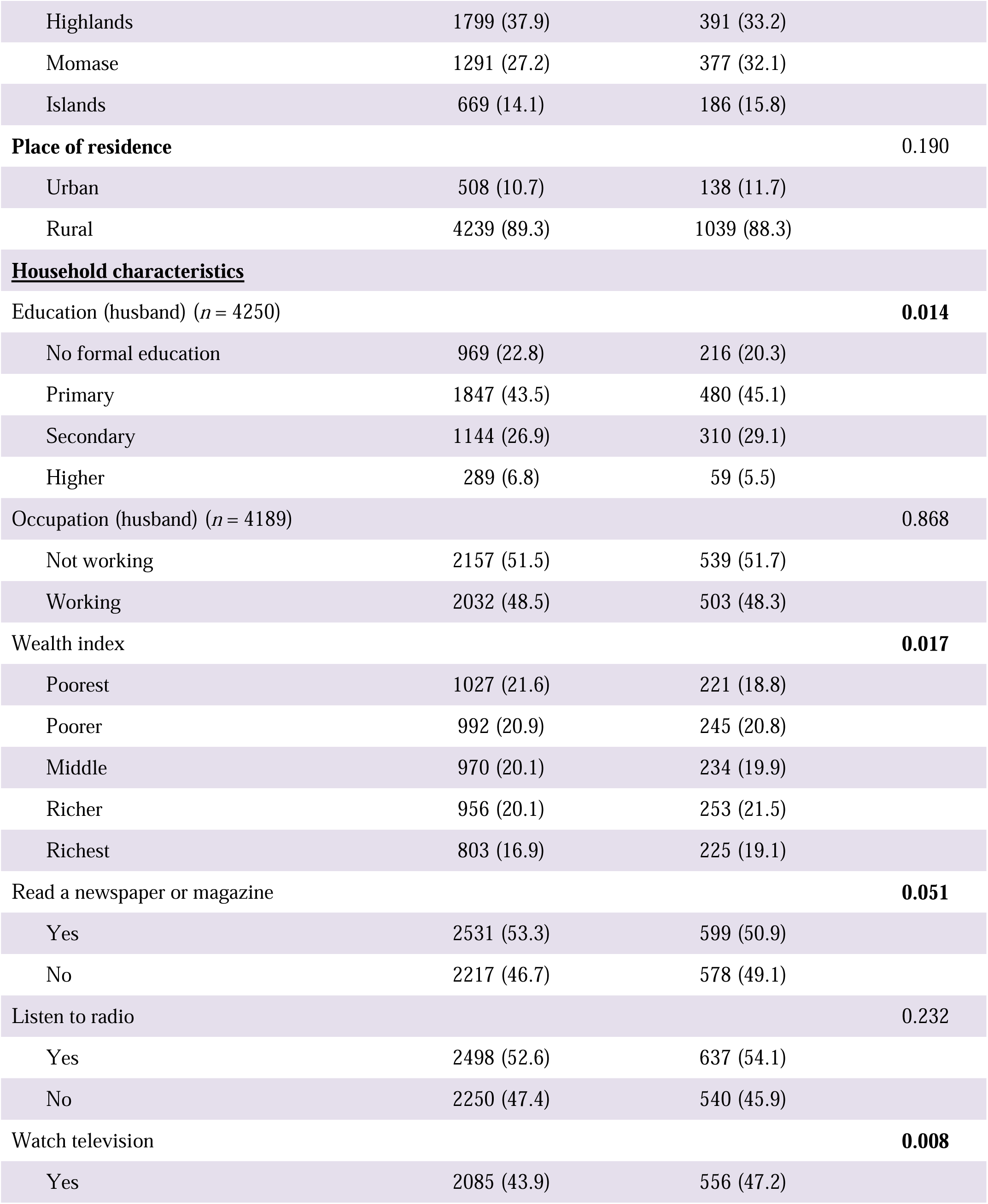
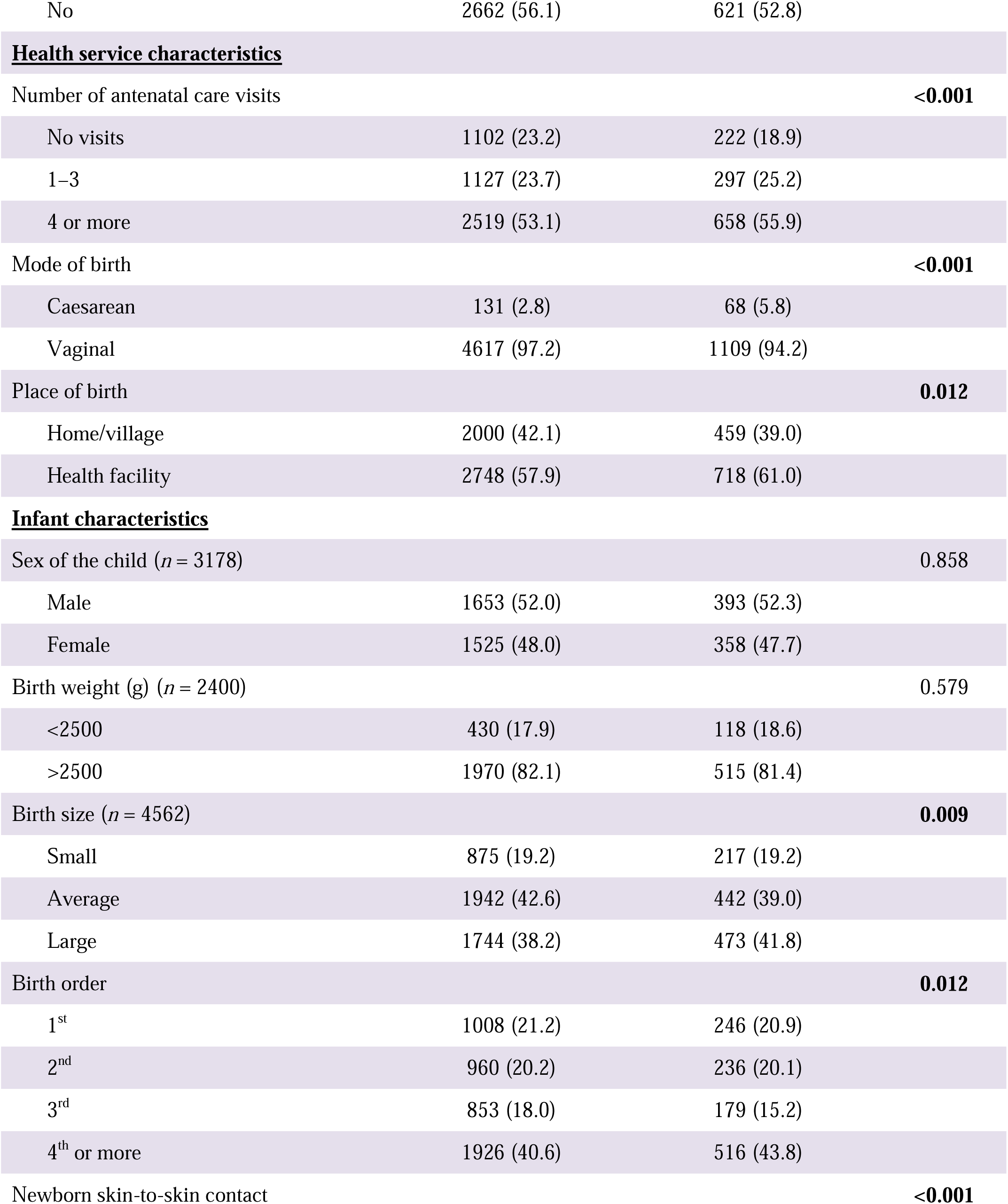
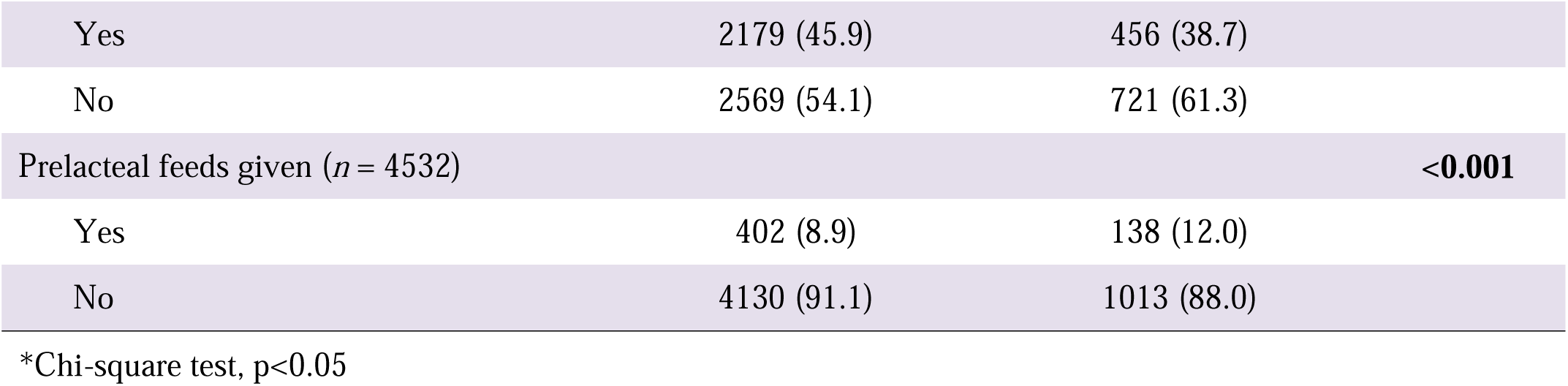
Characteristics of the participants.

### Factors associated with delayed initiation of breastfeeding

**Table 2** presents the multivariable analysis of determinants of delayed initiation of breastfeeding. Last pregnancy planned, reading a newspaper or magazine, watching television, region, mode of birth, place of birth, and newborn skin-to-skin contact immediately after birth were associated with delayed initiation of breastfeeding. In the multivariable analysis, the odds of delayed initiation of breastfeeding remained higher among women with an unplanned pregnancy (AOR 1.32; 95% CI: 1.03 to 1.68, p=0.030), those who had a caesarean section (AOR 3.16; 95% CI: 1.39 to 7.17, p=0.006), those who did not have initiate newborn skin-to-skin contact immediately after birth (AOR 1.83; 95% CI: 1.41 to 2.38, p<0.001), and those who watched television (AOR 1.39; 95% CI: 1.11 to 1.75, p=0.004), or were from the Momase region (AOR 1.31; 95% CI: 1.00 to 1.93). Conversely, women who read a newspaper or magazine (AOR 0.76; 95% CI: 0.61 to 0.95, p=0.018), were from the Southern (AOR 0.81; 95% CI: 0.56 to 1.15, p=0.041) and Highlands (AOR 0.86; 95% CI: 0.58 to 1.29, p=0.041) regions, and gave birth at home or in the village (AOR 0.69; 95% CI: 0.49 to 0.96, p=0.028), had lower odds of delaying initiation of breastfeeding. The study found no significant correlation between the number of antenatal care visits, birth size, birth order, prelacteal feeding and delayed initiation of breastfeeding.

**Table 2.**
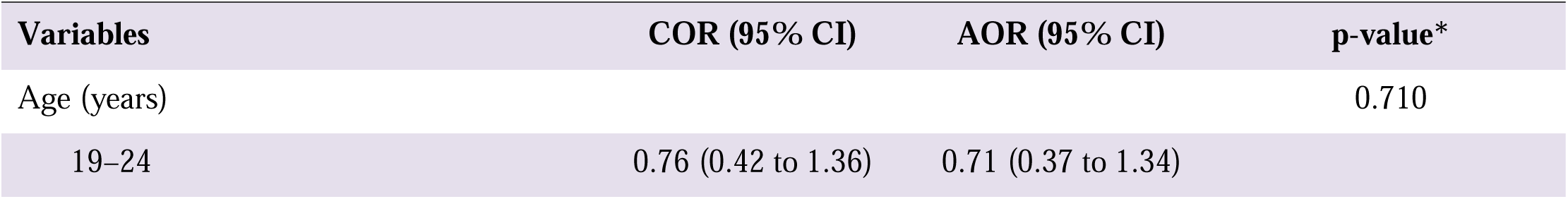
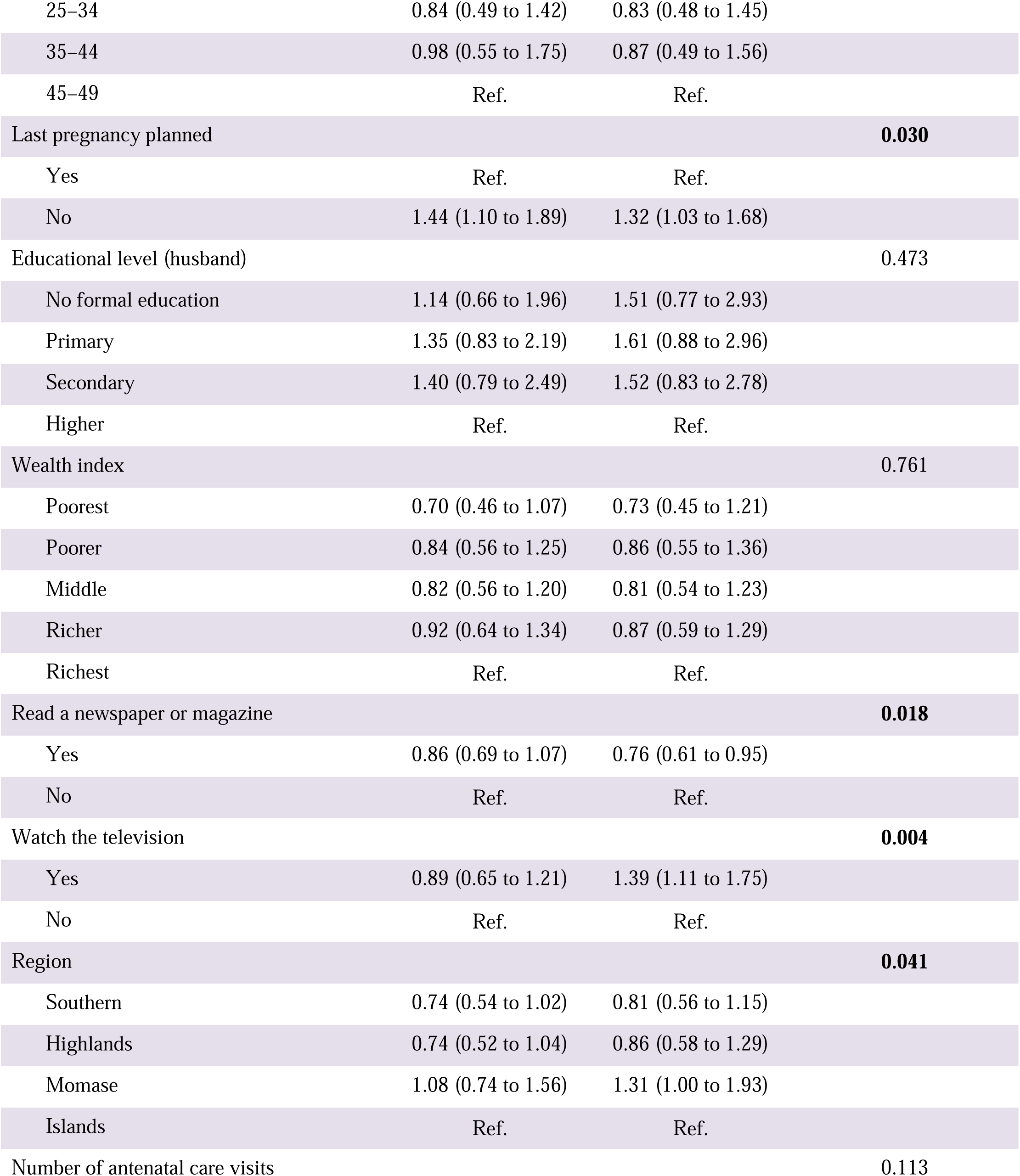
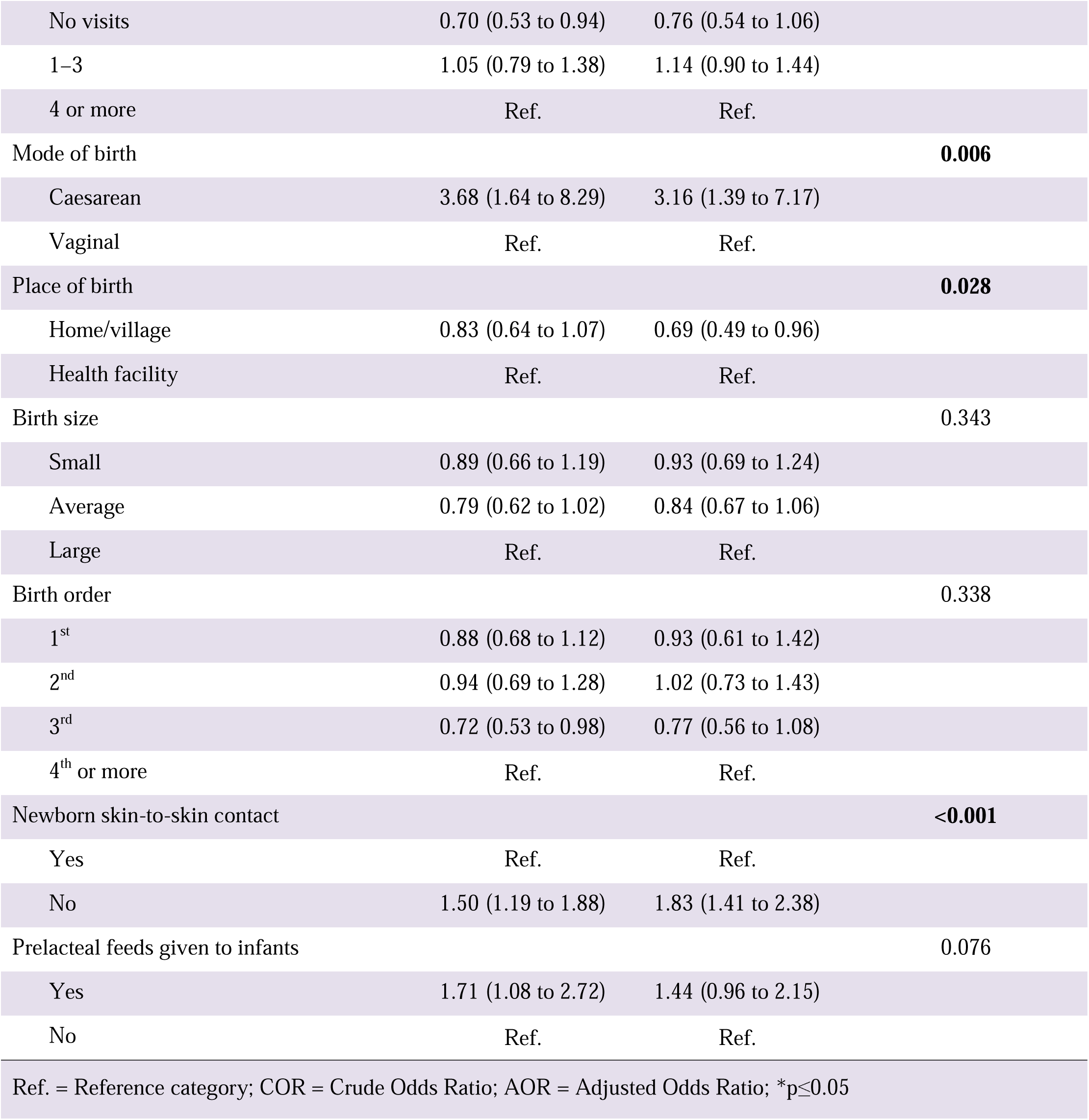
Multivariable analysis of factors associated with delayed initiation of breastfeeding in PNG.

| Variables | COR (95% CI) | AOR (95% CI) | p-value* |
| --- | --- | --- | --- |
| Age (years) |  |  | 0.710 |
| 19–24 | 0.76 (0.42 to 1.36) | 0.71 (0.37 to 1.34) |  |
| 25–34 | 0.84 (0.49 to 1.42) | 0.83 (0.48 to 1.45) |  |
| 35–44 | 0.98 (0.55 to 1.75) | 0.87 (0.49 to 1.56) |  |
| 45–49 | Ref. | Ref. |  |
| Last pregnancy planned |  |  | **0.030** |
| Yes | Ref. | Ref. |  |
| No | 1.44 (1.10 to 1.89) | 1.32 (1.03 to 1.68) |  |
| Educational level (husband) |  |  | 0.473 |
| No formal education | 1.14 (0.66 to 1.96) | 1.51 (0.77 to 2.93) |  |
| Primary | 1.35 (0.83 to 2.19) | 1.61 (0.88 to 2.96) |  |
| Secondary | 1.40 (0.79 to 2.49) | 1.52 (0.83 to 2.78) |  |
| Higher | Ref. | Ref. |  |
| Wealth index |  |  | 0.761 |
| Poorest | 0.70 (0.46 to 1.07) | 0.73 (0.45 to 1.21) |  |
| Poorer | 0.84 (0.56 to 1.25) | 0.86 (0.55 to 1.36) |  |
| Middle | 0.82 (0.56 to 1.20) | 0.81 (0.54 to 1.23) |  |
| Richer | 0.92 (0.64 to 1.34) | 0.87 (0.59 to 1.29) |  |
| Richest | Ref. | Ref. |  |
| Read a newspaper or magazine |  |  | **0.018** |
| Yes | 0.86 (0.69 to 1.07) | 0.76 (0.61 to 0.95) |  |
| No | Ref. | Ref. |  |
| Watch the television |  |  | **0.004** |
| Yes | 0.89 (0.65 to 1.21) | 1.39 (1.11 to 1.75) |  |
| No | Ref. | Ref. |  |
| Region |  |  | **0.041** |
| Southern | 0.74 (0.54 to 1.02) | 0.81 (0.56 to 1.15) |  |
| Highlands | 0.74 (0.52 to 1.04) | 0.86 (0.58 to 1.29) |  |
| Momase | 1.08 (0.74 to 1.56) | 1.31 (1.00 to 1.93) |  |
| Islands | Ref. | Ref. |  |
| Number of antenatal care visits |  |  | 0.113 |
| No visits | 0.70 (0.53 to 0.94) | 0.76 (0.54 to 1.06) |  |
| 1–3 | 1.05 (0.79 to 1.38) | 1.14 (0.90 to 1.44) |  |
| 4 or more | Ref. | Ref. |  |
| Mode of birth |  |  | **0.006** |
| Caesarean | 3.68 (1.64 to 8.29) | 3.16 (1.39 to 7.17) |  |
| Vaginal | Ref. | Ref. |  |
| Place of birth |  |  | **0.028** |
| Home/village | 0.83 (0.64 to 1.07) | 0.69 (0.49 to 0.96) |  |
| Health facility | Ref. | Ref. |  |
| Birth size |  |  | 0.343 |
| Small | 0.89 (0.66 to 1.19) | 0.93 (0.69 to 1.24) |  |
| Average | 0.79 (0.62 to 1.02) | 0.84 (0.67 to 1.06) |  |
| Large | Ref. | Ref. |  |
| Birth order |  |  | 0.338 |
| 1<sup>st</sup> | 0.88 (0.68 to 1.12) | 0.93 (0.61 to 1.42) |  |
| 2<sup>nd</sup> | 0.94 (0.69 to 1.28) | 1.02 (0.73 to 1.43) |  |
| 3<sup>rd</sup> | 0.72 (0.53 to 0.98) | 0.77 (0.56 to 1.08) |  |
| 4<sup>th</sup> or more | Ref. | Ref. |  |
| Newborn skin-to-skin contact |  |  | **<0.001** |
| Yes | Ref. | Ref. |  |
| No | 1.50 (1.19 to 1.88) | 1.83 (1.41 to 2.38) |  |
| Prelacteal feeds given to infants |  |  | 0.076 |
| Yes | 1.71 (1.08 to 2.72) | 1.44 (0.96 to 2.15) |  |
| No | Ref. | Ref. |  |
| Ref. = Reference category; COR = Crude Odds Ratio; AOR = Adjusted Odds Ratio; *p≤0.05 |  |  |  |

## Discussion

This study sought to determine the prevalence of delayed initiation of breastfeeding in PNG and to examine the association between household, maternal, child, and health services determinants. Women who watched television, were from the Momase region, had not planned their last pregnancies, had a caesarean section, and had no newborn skin-to-skin contact immediately after birth were more likely to experience a delay in initiating breastfeeding. The findings in this study underscore the demand for tailored program interventions to improve breastfeeding initiation practices, particularly among rural women.

While timely breastfeeding initiation is crucial for the growth and survival of infants, the practice remains suboptimal in PNG. In this study, about one-quarter (24.7%) of women have delayed breastfeeding initiation. This result was lower than the findings from an earlier study in PNG,^12^ including those in Bangladesh,^4^ South Sudan,^21^ and Uganda.^22^ The variation in prevalence may be due to sample size, maternal socio-demographic factors (i.e., socio-economic status, educational status, social norms and beliefs concerning breastfeeding practice), and health service utilisation.

Findings from this study revealed that women with unplanned pregnancies were more likely to delay initiation of breastfeeding. This is in agreement with studies done in sub-Saharan Africa.^23^ Women who have unplanned pregnancies likely underutilise maternal health services, such as family planning and antenatal care, which often include appropriate information and education on breastfeeding for pregnant women.^24,25^ Despite this finding, the increased odds of delayed initiation of breastfeeding with unplanned pregnancies among women in PNG are an observation that would require further qualitative inquiry.

Caesarean section was positively associated with delayed initiation of breastfeeding in this study, consistent with several studies conducted elsewhere.^26–28^ Women who underwent caesarean section were three times more likely to have a delayed initiation of breastfeeding. This could be explained by the hospital practice of separating newborns from their mothers after the procedure.^28^ In addition, maternal exhaustion and the effects of anaesthesia following a caesarean section may impede early breastfeeding initiation.^22,29^ While maternal recovery from this procedure generally takes some time, immediate breastfeeding is required once the woman regains consciousness and alertness.^30,31^

Interestingly, women who gave birth at home or in the village had lower odds of delayed initiation of breastfeeding compared with those who gave birth in a health facility. This was similar to findings from several LMICs, ^32^ indicating that home-birthed infants were more likely to be breastfed in the first hour of birth. The type and level of support from skilled birth attendants, family members, particularly partners/husbands, and a familiar home birthing environment may reduce stress, leading to intervention-free birth, and consequently influence breastfeeding outcomes.^32,33^ Evidence from studies conducted in PNG has demonstrated an inverse association between social and family support that adversely affects breastfeeding initiation for mothers who had given birth at home or in the village.^13–15^ However, it is unclear what underlies the observed association between delayed initiation of breastfeeding and home/village birth in the current study. Furthermore, the association between home/village birth and breastfeeding initiation is improbable to be directly causal, which will require further research to confirm the factors driving the observed differences.

From the results, poor newborn skin-to-skin contact immediately after birth was significantly associated with delayed initiation of breastfeeding. Women who did not initiate skin-to-skin contact immediately after birth with their newborns were more likely to delay initiation of breastfeeding. They may be unaware of the importance of skin-to-skin contact as a means of promoting warmth and thermoregulation for their newborns, thus their lower odds of embracing and practising it.^12,34^ The lack of information may therefore account for the delayed initiation of breastfeeding. Certain socio-cultural norms and beliefs may disapprove of these women for practising newborn skin-to-skin contact.^13,15,34^ Furthermore, poor knowledge on the part of health workers, shortages of health workers, poor newborn care, and limited time can impede the practice of newborn skin-to-skin contact.^34,35^ Qualitative inquiry is necessary to investigate the knowledge, attitudes, and practices regarding breastfeeding initiation among health workers and mothers, particularly those in rural settings.

Women who were exposed to mass media in this study, particularly television, were more likely to delay initiation of breastfeeding, consistent with studies done in sub-Saharan Africa.^23,36^ Social marketing and advertising of infant formula feedings, milk substitutes, teats, and bottles on various media platforms have been shown to affect breastfeeding initiation.^37,38^ In contrast, this study further revealed that women who read newspapers or magazines had lower odds of delaying breastfeeding initiation. Print media (i.e., newspapers or magazines) are available and accessible to the public compared to television, which could explain the reason for this finding. Access to print media improves information and health literacy on breastfeeding and its benefits and fosters positive behavioural beliefs about breastfeeding which have been reported as important determinants of optimal breastfeeding practice.^39–41^ Print media can be an influential tool for breastfeeding communication and promotion, for persuading breastfeeding behaviours, creating positive social norms, and garnering support from stakeholders and policymakers.^40^ However, limited information exists on how mass media coverage influences breastfeeding practice in PNG’s context. Further qualitative inquiry for a deeper understanding of mass media coverage of breastfeeding practices is necessary.

Regarding the region, delayed initiation of breastfeeding was not common among women from the Highlands and Southern regions. This finding is in agreement with a similar study conducted in PNG.^12^ Expansion of access to healthcare services and ongoing economic developments in these two regions are possible explanations for this observation. On the contrary, the odds of delayed breastfeeding initiation remained higher among women from the Momase region compared to women in the Island region. This is likely attributed to the lack of qualified health workers with adequate resources in this region of the country, impeding access to maternal, newborn, and child health services as reported in similar studies.^12,42^ This can also be elucidated in terms of the socioeconomic condition, frequently characterized by poor household wealth and low education levels, including accessibility, attitudes to antenatal care, and interpersonal issues affecting women from accessing the services provided by health workers in this region.^42,43^ Most of the provinces in the Momase region are relatively economically unstable and geographically remote, which makes healthcare, services problematic.^43^ The regional variations relating to breastfeeding initiation could be attributed to poor healthcare service and resource distribution. This is also consistent with previous studies, which found that variations exist in various regions and the utilisation of antenatal care services.^42,43^ Thus, mobile and outreach programs should be strengthened to reach women, especially in rural areas and the disadvantaged.

This study has several strengths. It investigated household, maternal, and child factors and healthcare utilisation with delayed initiation of breastfeeding using a nationally representative survey. In addition, appropriate analysis techniques such as weighting and complex samples analysis were used to ensure that the results were representative of women in PNG. The two-stage sampling approach ensured that no selection bias could influence the results of this study. However, the study has limitations and should be interpreted with caution. As it was a cross-sectional study, the analysis could not determine a cause-and-effect relationship between breastfeeding initiation and independent variables. The role of health workers and skilled birth attendants (e.g., midwives or nurses) regarding breastfeeding was not captured in the dataset; therefore, the extent of breastfeeding counselling and support provided during antenatal and early postpartum periods could not be established. Since the outcome of the study was assessed based on the women’s responses regarding the timing of breastfeeding initiation, results may have been influenced by self-report and social desirability biases. Furthermore, the DHS did not collect some information, such as maternal beliefs and knowledge about breastfeeding, so there may be residual confounding.

## Conclusion

Early initiation of breastfeeding within one hour after birth is beneficial in reducing neonatal mortality and preventing other neonatal illnesses. This study found that one in four women delayed early breastfeeding initiation. Factors significantly associated with delayed initiation of breastfeeding include unplanned pregnancies, media exposure, region, caesarean section, and poor newborn skin-to-skin contact immediately after birth. Interventions to promote optimal breastfeeding require a multi-sectoral approach, as well as bolstering health workers’ capacity to encourage and support early initiation of breastfeeding during the antenatal and early postnatal periods. This could increase breastfeeding initiation rates at birth and prevent adverse neonatal outcomes in PNG.

## Data Availability Statement

The data underlying the results presented in the study are available from the DHS Program: https://dhsprogram.com/methodology/survey/survey-display-499.cfm.

## Acknowledgements

The authors acknowledge the DHS Program for making the datasets available for this analysis and the women who participated in the surveys.

## Contributors

MM conceived and designed the study protocol. MM prepared the data, performed the statistical data analysis and produced the tables and figures. MM, FP and LMV interpreted the data. MM drafted the manuscript. FP and LMV critically revised and edited the manuscript for important intellectual content. All authors read and approved the final manuscript. MM is the guarantor and accepts full responsibility for the work.

## Funding

The authors have not declared a specific grant for this research from any funding agency in the public, commercial or not-for-profit sectors.

## Competing interests

None declared.

## Patient consent for publication

Not applicable.

## Ethics approval

The 2016–2018 PNGDHS protocol was reviewed and approved by the Institutional Review Board of Inner-City Fund (ICF) International (530 Gaither Road, Suite 500 Rockville, MD 20850, USA). The dataset was accessed through the DHS program website (https://www.dhsprogram.com/data) after the user’s agreement and approval for use were completed. The data used in this study were de-identified to ensure anonymity. Informed consent was obtained from all participants before the survey was conducted. Therefore, no additional ethical approval was sought for this study.

## Provenance and peer review

Not commissioned; externally peer-reviewed

## Notes

### Competing Interest Statement

The authors have declared no competing interest.

### Funding Statement

This study did not receive any funding.

### Author Declarations

The study used (or will use) ONLY openly available human data that were originally located at: https://dhsprogram.com/methodology/survey/survey-display-499.cfm.

## References

1. NEOVITA Study Group. Timing of initiation, patterns of breastfeeding, and infant survival: prospective analysis of pooled data from three randomised trials. The Lancet Global Health. 2016 Apr 1;4(4):e266–75.

2. Victora CG, Bahl R, Barros AJD, França GVA, Horton S, Krasevec J, et al. Breastfeeding in the 21st century: epidemiology, mechanisms, and lifelong effect. The Lancet. 2016 Jan 30;387(10017):475–90.

3. WHO/UNICEF. Capture the Moment – Early initiation of breastfeeding: The best start for every newborn [Internet]. 2018. Available from: https://data.unicef.org/resources/capture-the-moment/

4. Raihana S, Alam A, Huda TM, Dibley MJ. Factors associated with delayed initiation of breastfeeding in health facilities: secondary analysis of Bangladesh demographic and health survey 2014. International Breastfeeding Journal. 2021 Jan 22;16(1):14.

5. Abedi P, Jahanfar S, Namvar F, Lee J. Breastfeeding or nipple stimulation for reducing postpartum haemorrhage in the third stage of labour. Cochrane Database of Systematic Reviews [Internet]. 2016;(1). Available from: 10.1002/14651858.CD010845.pub2

6. Duale A, Singh P, Al Khodor S. Breast milk: a meal worth having. Frontiers in Nutrition. 2022;8:800927.

7. Khan J, Vesel L, Bahl R, Martines JC. Timing of Breastfeeding Initiation and Exclusivity of Breastfeeding During the First Month of Life: Effects on Neonatal Mortality and Morbidity—A Systematic Review and Meta-analysis. Maternal and Child Health Journal. 2015 Mar 1;19(3):468–79.

8. Morris A. Breastfeeding reduces risk of type 2 diabetes mellitus. Nature Reviews Endocrinology. 2018 Mar 1;14(3):128–128.

9. Jelly P, Choudhary S. Breastfeeding and breast cancer: A risk reduction strategy. IP Int J Med Paediatr Oncol. 2019;5(2):47–50.

10. Bruno Tongun J, Sebit MB, Mukunya D, Ndeezi G, Nankabirwa V, Tylleskar T, et al. Factors associated with delayed initiation of breastfeeding: a cross-sectional study in South Sudan. International Breastfeeding Journal. 2018 Jul 5;13(1):28.

11. Smith ER, Hurt L, Chowdhury R, Sinha B, Fawzi W, Edmond KM, et al. Delayed breastfeeding initiation and infant survival: A systematic review and meta-analysis. PLOS ONE. 2017 Jul 26;12(7):e0180722.

12. Seidu AA, Ahinkorah BO, Agbaglo E, Dadzie LK, Tetteh JK, Ameyaw EK, et al. Determinants of early initiation of breastfeeding in Papua New Guinea: a population-based study using the 2016-2018 demographic and health survey data. Archives of Public Health. 2020 Nov 23;78(1):124.

13. Maviso MK, Kaforau LM, Hastie C. Influence of grandmothers on breastfeeding practices in a rural community in Papua New Guinea: A critical discourse analysis of first- time mothers’ perspectives. Women and Birth. 2023;36(2):e263–9.

14. Maviso MK, Ferguson B, Kaforau LM, Capper T. A qualitative descriptive inquiry into factors influencing early weaning and breastfeeding duration among first-time mothers in Papua New Guinea’s rural eastern highlands. Women and Birth. 2022;35(1):e68–74.

15. Terry B, Kebio L, Kuzma J. Knowledge, attitude and practice of mothers towards breastfeeding in rural Papua New Guinea: a mixed method study. Papua New Guinea Medical Journal. 2015;58(1/4):22.

16. Kuzma J. Knowledge, attitude and practice related to infant feeding among women in rural Papua New Guinea: a descriptive, mixed method study. International Breastfeeding Journal. 2013 Nov 21;8(1):16.

17. National Department of Health. Infant & Young Child Feeding Policy 2014. Port Moresby, Papua New Guinea: National Department of Health; 2014.

18. National Statistical Office (NSO) [Papua New Guinea], ICF. Papua New Guinea Demographic and Health Survey 2016-18 [Internet]. Port Moresby, Papua New Guinea, and Rockville, Maryland, USA: NSO and ICF; 2019. Available from: https://www.dhsprogram.com/pubs/pdf/FR364/FR364.pdf

19. Kambale RM, Buliga JB, Isia NF, Muhimuzi AN, Battisti O, Mungo BM. Delayed initiation of breastfeeding in Bukavu, South Kivu, eastern Democratic Republic of the Congo: a cross-sectional study. International Breastfeeding Journal. 2018 Feb 13;13(1):6.

20. Archer KJ, Lemeshow S, Hosmer DW. Goodness-of-fit tests for logistic regression models when data are collected using a complex sampling design. Computational Statistics & Data Analysis. 2007 May 15;51(9):4450–64.

21. Bruno Tongun J, Sebit MB, Mukunya D, Ndeezi G, Nankabirwa V, Tylleskar T, et al. Factors associated with delayed initiation of breastfeeding: a cross-sectional study in South Sudan. International Breastfeeding Journal. 2018 Jul 5;13(1):28.

22. Mukunya D, Tumwine JK, Nankabirwa V, Ndeezi G, Odongo I, Tumuhamye J, et al. Factors associated with delayed initiation of breastfeeding: a survey in Northern Uganda. Global Health Action. 2017 Jan 1;10(1):1410975.

23. Teshale AB, Tesema GA. Timely initiation of breastfeeding and associated factors among mothers having children less than two years of age in sub-Saharan Africa: A multilevel analysis using recent Demographic and Health Surveys data. PLOS ONE. 2021 Mar 23;16(3):e0248976.

24. Bekele H, Dheressa M, Mengistie B, Sintayehu Y, Fekadu G. Unintended Pregnancy and Associated Factors among Pregnant Women Attending Antenatal Care at Bako Tibe District Public Health Facility, Oromia Region, Ethiopia. Balayla J, editor. Journal of Pregnancy. 2020 Mar 19;2020:3179193.

25. Hromi-Fiedler AJ, Pérez-Escamilla R. Unintended pregnancies are associated with less likelihood of prolonged breast-feeding: an analysis of 18 Demographic and Health Surveys. Public Health Nutrition. 2007/01/02 ed. 2006;9(3):306–12.

26. Sharma M, Anand A, Goswami I, Pradhan MR. Factors associated with delayed initiation and non-exclusive breastfeeding among children in India: evidence from national family health survey 2019-21. International Breastfeeding Journal. 2023 Jun 6;18(1):28.

27. Kusasira L, Mukunya D, Obakiro S, Kenedy K, Rebecca N, Ssenyonga L, et al. Prevalence and predictors of delayed initiation of breastfeeding among postnatal women at a tertiary hospital in Eastern Uganda: a cross-sectional study. Archives of Public Health. 2023 Apr 14;81(1):56.

28. Tongun JB, Sebit MB, Ndeezi G, Mukunya D, Tylleskar T, Tumwine JK. Prevalence and determinants of pre-lacteal feeding in South Sudan: a community-based survey. Global Health Action. 2018 Jan 1;11(1):1523304.

29. Hobbs AJ, Mannion CA, McDonald SW, Brockway M, Tough SC. The impact of caesarean section on breastfeeding initiation, duration and difficulties in the first four months postpartum. BMC Pregnancy and Childbirth. 2016 Apr 26;16(1):90.

30. Cobb B, Liu R, Valentine E, Onuoha O. Breastfeeding after Anesthesia: A Review for Anesthesia Providers Regarding the Transfer of Medications into Breast Milk. Transl Perioper Pain Med. 2015;1(2):1–7.

31. Mitchell J, Jones W, Winkley E, Kinsella SM. Guideline on anaesthesia and sedation in breastfeeding women 2020. Anaesthesia. 2020 Nov 1;75(11):1482–93.

32. Oakley L, Benova L, Macleod D, Lynch CA, Campbell OMR. Early breastfeeding practices: Descriptive analysis of recent Demographic and Health Surveys. Maternal & Child Nutrition. 2018 Apr 1;14(2):e12535.

33. Oyedele OK. Correlates of non-institutional delivery to delayed initiation of breastfeeding in Nigeria: logit-decomposition and subnational analysis of population-based survey. Journal of Health, Population and Nutrition. 2023 Nov 6;42(1):121.

34. Aboagye RG, Okyere J, Dowou RK, Adzigbli LA, Tackie V, Ahinkorah BO, et al. Prevalence and predictors of mother and newborn skin-to-skin contact at birth in Papua New Guinea. BMJ Open. 2022 Sep 5;12(9):e062422.

35. Wilson AN, Melepia P, Suruka R, Hezeri P, Kabiu D, Babona D, et al. Quality newborn care in East New Britain, Papua New Guinea: measuring early newborn care practices and identifying opportunities for improvement. BMC Pregnancy and Childbirth. 2022 Jun 1;22(1):462.

36. Appiah F, Ahinkorah BO, Budu E, Oduro JK, Sambah F, Baatiema L, et al. Maternal and child factors associated with timely initiation of breastfeeding in sub-Saharan Africa. International Breastfeeding Journal. 2021 Jul 19;16(1):55.

37. Mishel Unar-Munguía, Andrea Santos-Guzmán, Pedro Javier Mota-Castillo, Marena Ceballos-Rasgado, Lizbeth Tolentino-Mayo, Matthias Sachse Aguilera, et al. Digital marketing of formula and baby food negatively influences breast feeding and complementary feeding: a cross-sectional study and video recording of parental exposure in Mexico. BMJ Global Health. 2022 Nov 1;7(11):e009904.

38. Henjum S, Mathisen R, Nguyen TT, Phan LTH, Ribe LO, Vinje KH. Media audit reveals inappropriate promotion of products under the scope of the International Code of Marketing of Breast-milk Substitutes in South-East Asia. Public Health Nutrition. 2017/03/15 ed. 2017;20(8):1333–42.

39. Frerichs L, Andsager JL, Campo S, Aquilino M, Dyer CS. Framing Breastfeeding and Formula-Feeding Messages in Popular U.S. Magazines. Women & Health. 2006 Nov 28;44(1):95–118.

40. Merritt R, Eida T, Safon C, Kendall S. Print media coverage of breastfeeding in Great Britain: Positive or negative? Maternal & Child Nutrition. 2023 Jan 1;19(S1):e13458.

41. Hitt R, Zhuang J, Anderson J. Media Presentation of Breastfeeding Beliefs in Newspapers. Health Communication. 2018 Oct 3;33(10):1293–301.

42. Seidu AA, Agbaglo E, Dadzie LK, Ahinkorah BO, Ameyaw EK, Tetteh JK. Individual and contextual factors associated with barriers to accessing healthcare among women in Papua New Guinea: insights from a nationwide demographic and health survey. International Health. 2021 Nov 1;13(6):573–85.

43. Li Y, Li H, Jiang Y. Factors influencing maternal healthcare utilization in Papua New Guinea: Andersen’s behaviour model. BMC Women’s Health. 2023 Oct 21;23(1):544.

44. Raihana S, Alam A, Chad N, Huda TM, Dibley MJ. Delayed Initiation of Breastfeeding and Role of Mode and Place of Childbirth: Evidence from Health Surveys in 58 Low- and Middle- Income Countries (2012–2017). International Journal of Environmental Research and Public Health. 2021;18(11).

45. Hector D, King L, Webb K, Heywood P. Factors affecting breastfeeding practices. Applying a conceptual framework. NSW Public Health Bull. 2005;16(4):52–5.

